# Prodromal Progressive Supranuclear Palsy – insights from the UK Biobank

**DOI:** 10.1101/2021.05.06.21256759

**Authors:** Duncan Street, David Whiteside, Timothy Rittman, James B Rowe

**Author notes:** **Corresponding author:** Professor James B. Rowe, Department of Clinical Neurosciences, University of Cambridge, Herchel Smith Building, Forvie Site, Robinson Way, Cambridge Biomedical Campus, Cambridge, CB2 0SZ. **Financial Disclosure:** Dr Street, Dr Whiteside and Dr Rittman report no disclosures. Professor Rowe serves as editor to *Brain* and is a non-remunerated trustee of the Guarantors of Brain and the PSP Association (UK). He provides consultancy to Asceneuron, Biogen, UCB, SVHealth and Wave, and has research grants from AZ-Medimmune, Janssen, Lilly as industry partners in the Dementias Platform UK. **Funding:** This study was co-funded by the National Institute for Health Research (NIHR) Biomedical Research Centre at Cambridge University Hospitals NHS Foundation Trust and the University of Cambridge (BRC-1215-20014) and the Cambridge Brain Bank; the Holt Fellowship (RG86564); the Medical Research Council (MR/P01271X/1; SUAG051/G101400 and SUAG004/051/RG91365); and the Cambridge Centre for Parkinson-plus (RG95450). The views expressed are those of the authors and not necessarily those of the NIHR or the Department of Health and Social Care.

## Abstract

**Background:** Prodromal Parkinson’s Disease is well described but prodromal Progressive Supranuclear Palsy (PSP) is much less understood. The diagnosis of PSP is typically delayed by an average of three years after symptom onset. Understanding the changes that occur in the prodromal and pre-diagnostic period will aid earlier diagnosis, clarify the natural history, and aid the design of early disease-modifying therapy trials.

**Objectives:** To determine motor and cognitive markers of prodromal PSP, with Parkinson’s disease as a comparator condition, in a large prospective cohort.

**Methods:** Baseline UK Biobank data from 502,504 individuals were collected between 2006 and 2010. Subsequent PSP and Parkinson’s disease cases were identified from primary and secondary care electronic health records’ diagnostic coding data and death registry, with 5,404 matched controls.

**Results:** 176 PSP cases (time to diagnosis 7.8±2.8 years) and 2,526 Parkinson’s disease cases (time to diagnosis 7.8±2.9 years) were identified. At baseline, those later diagnosed with PSP had slower reaction times, weaker hand grip, lower fluid intelligence, poorer prospective memory, worse self-rated health score and lower digit recall than controls. They had higher mortality than both Parkinson’s disease and control groups.

**Conclusions:** Motor slowing, cognitive dysfunction, and postural instability are clinical diagnostic features of PSP and are typically symptomatic three years before diagnosis. However, objective markers of these features are evident over seven years before diagnosis. This suggests a long prodromal course in PSP with subtle changes in motor and cognitive function.

## Introduction

Progressive Supranuclear Palsy (PSP) is a devastating and rapidly progressive disorder with a poor prognosis once diagnosed, compounded by a typical delay between symptom onset and diagnosis of over three years.^1,2^ In contrast to Parkinson’s disease and genetic forms of dementia (e.g. Alzheimer’s disease, Frontotemporal dementia, Huntington’s disease), little is known about this pre-diagnostic or prodromal period. Community-based ageing and brain bank donation studies have identified neuropathological features of PSP in people who were undiagnosed in life.^3–6^ However, progress towards better definition of the prodromal phase has been hampered by the scarcity of PSP, the lack of a clear genetic aetiology and small, frequently underpowered retrospective case-control studies presenting a high risk of recall and selection bias depending on recruitment methodologies, and differences between clinical diagnosis and brain bank criteria.^7^

The UK Biobank provides an important and less biased opportunity to study the prodromal phase in PSP. Over 500,000 middle-aged individuals were recruited between 2006 to 2010. Baseline assessment consisted of demographics, medical history, cognitive function, and physical measures. Continual supplementation of medical history is provided through ongoing access to primary and secondary care electronic diagnostic coding records and death registry data. As such, this dataset offers a unique window into subtle changes during the pre-diagnostic and pre-symptomatic phases of PSP.

As a disease control, we compared the prodromal features of PSP with the prodromal features of Parkinson’s disease. The prodromal phase of Parkinson’s disease has formal diagnostic criteria^8,9^ and may last between 5 and 20 years prior to diagnosis.^10,11^ The current criteria^9^ recognise cognitive deficits and abnormal quantitative motor test performance as useful prodromal markers while subjective self-reported symptoms showed promise. Despite differing pathological substrates and pathophysiological mechanisms, the shared clinical features, classification, and referral pathways offer enough similarity to provide a valuable benchmark against which to compare the prodromal features of PSP.

By analogy with common and genetic neurodegenerative disorders,^12–16^ we proposed the existence of a long prodromal period for PSP, predicting subtle cognitive and motor abnormalities on objective testing even in the absence of symptoms or diagnosis. We test this hypothesis using the UK Biobank longitudinal data. We use the term ‘prodromal’ to refer to a stage of mild symptomatic manifestation and the term ‘pre-symptomatic’ to refer to a stage before symptoms are present but when changes may be detected on objective tests or examination.

## Methods

### Participants

The UK Biobank (UKB) is a large prospective, population-based cohort study of over 500,000 individuals aged 40-69 years old recruited between 2006 and 2010 from one of 22 UK centres.^17^ The UKB was established to investigate genetic, health and lifestyle determinants of disease in middle-aged and older adults and baseline assessment included questionnaires, face-to-face interviews (including tests of cognitive function), physical measures (e.g. hand grip strength, reaction time), blood sampling, genetic analysis and imaging. Cognitive function assessment included tests of prospective and retrospective memory and fluid intelligence. Participants gave informed consent to allow ongoing access to their medical records to include primary and secondary care diagnostic coding data (according to ICD-10 classification), and death registry. Data were accessed through an approved application to the UKB (Application ID: 46620).

PSP and Parkinson’s disease cases were identified through patient self-report at assessment, primary and secondary care diagnostic coding data and death registry and were matched with age and gender-similar controls (2:1). The date of first coding of a diagnosis was used as a proxy for the date of diagnosis and when a coding diagnosis was recorded through multiple sources the earliest date was used. Those with a known diagnosis at study baseline were excluded. The census date was set as 01/11/2020.

### Statistical analysis

R studio (version 4.0.3, R Core Team, 2020) was used for analysis of demographic and clinical data. The *survival*^18^ and *survminer*^19^ packages in R studio were used to construct Cox Survival regression models. The *MatchIt*^20^ package was used for case-control age and sex matching. The Shapiro-Wilk test was used to assess for normality of data distribution. Comparison of means was performed using the independent samples t-test when normally distributed and the Kruskal-Wallis test when not normally distributed. Mean values of continuous data are expressed with their associated standard deviations. Categorical data were compared using the Chi Square test. Variables within the survival model are detailed along with their associated hazard ratios (HR) and 95% confidence intervals (CI). The Log-Rank test was used to compare survival curves. For all analyses, significance level was adjusted by Bonferroni’s correction and a p<.05 was considered significant.

## Results

The UK Biobank sample comprised 502,504 individuals. Mean age at baseline assessment was 57.1±8.1 years. Forty-six percent were male. Six percent (32,518/502,504) had died by the census date.

Demographic data for people later developing PSP and Parkinson’s disease, and matched controls, are presented in Table 1. One hundred and seventy-six PSP cases and 2,526 Parkinson’s disease cases were identified. Primary care records were the source for the majority of initial Parkinson’s disease (1284/2526, 51%) and PSP (85/176, 48%) diagnostic coding. Fifty-five PSP cases (55/176, 31%) were initially coded as Parkinson’s disease with a mean time to revision of diagnosis of approximately 2 years. Age at baseline assessment, age at diagnosis and years from baseline assessment to diagnosis were similar between groups.

**Table 1:** Demographics, mortality and baseline quantitative test performance by diagnostic group

|  | Control | PD | PSP | Control vs PD | Control vs PSP | PD vs PSP |
| --- | --- | --- | --- | --- | --- | --- |
|  | *N=5404* | *N=2526* | *N=176* | *p-value* | *p-value* | *p-value* |
| **Demographics** |  |  |  |  |  |  |
| Source of diagnostic coding: |  |  |  | - | - | - |
| Death registry | - | 26 (1.03%) | 12 (6.82%) |  |  |  |
| Primary care | - | 1284 (50.8%) | 85 (48.3%) |  |  |  |
| Secondary care | - | 1216 (48.1%) | 79 (44.9%) |  |  |  |
| Age at baseline | 63.7 (5.27) | 63.7 (5.25) | 63.8 (5.66) | 0.926 | 0.802 | 0.802 |
| Sex, Male/Female (%) | 3346/2058<br>(62/38) | 1566/960<br>(62/38) | 105/71<br>(60/40) | 0.967 | 0.598 | 0.592 |
| Age at diagnosis | - | 71.5 (5.97) | 71.6 (6.35) | - | - | 0.593 |
| Time to diagnosis (years) | - | 7.80 (2.94) | 7.79 (2.76) | - | - | 0.748 |
| **Mortality** |  |  |  |  |  |  |
| Number dead (%) | 385 (7) | 712 (28) | 110 (62) | <0.001 | <0.001 | <0.001 |
| Age at death | 73.7 (4.83) | 74.4 (4.83) | 72.4 (6.25) | 0.011 | 0.116 | 0.003 |
| Assessment to death (years) | 7.39 (3.15) | 9.31 (2.36) | 8.68 (2.03) | <0.001 | 0.001 | 0.001 |
| Diagnosis to death (years) | - | 2.42 (2.46) | 1.60 (1.57) | - | - | 0.006 |
| **Quantitative Test parameters** |  |  |  |  |  |  |
| Weight (kg) | 78.8 (14.5) | 80.3 (15.4) | 82.4 (14.7) | 0.001 | 0.003 | 0.069 |
| Reaction time (ms) | 574 (117) | 596 (132) | 618 (158) | <0.001 | 0.001 | 0.147 |
| Left hand grip strength (kg) | 31.0 (11.0) | 28.8 (11.1) | 29.4 (10.6) | <0.001 | 0.053 | 0.619 |
| Right hand grip strength (kg) | 33.0 (11.0) | 31.0 (11.2) | 31.0 (10.3) | <0.001 | 0.014 | 0.839 |
| Pair matching (errors) | 4.38 (3.55) | 4.59 (3.62) | 4.77 (3.95) | 0.220 | 0.492 | 0.879 |
| Fluid intelligence score | 6.15 (2.09) | 5.70 (2.14) | 5.10 (2.12) | <0.001 | <0.001 | 0.029 |
| Digit recall score | 6.62 (1.75) | 6.18 (1.84) | 5.71 (1.98) | 0.001 | 0.027 | 0.286 |

PD = Parkinson's disease, PSP = Progressive Supranuclear palsy

Mortality data are presented in Table 1 and corresponding Kaplan-Meier survival curves are displayed in Figure 1. Significantly more of the PSP group had died by census date versus the Parkinson’s disease group. Both the PSP group and the Parkinson’s disease group demonstrated higher rates of mortality than age and gender matched controls. Compared to the Parkinson’s disease group, the PSP group was younger at death, had a shorter time from assessment to death and a shorter time from diagnosis to death.

**Figure 1:**
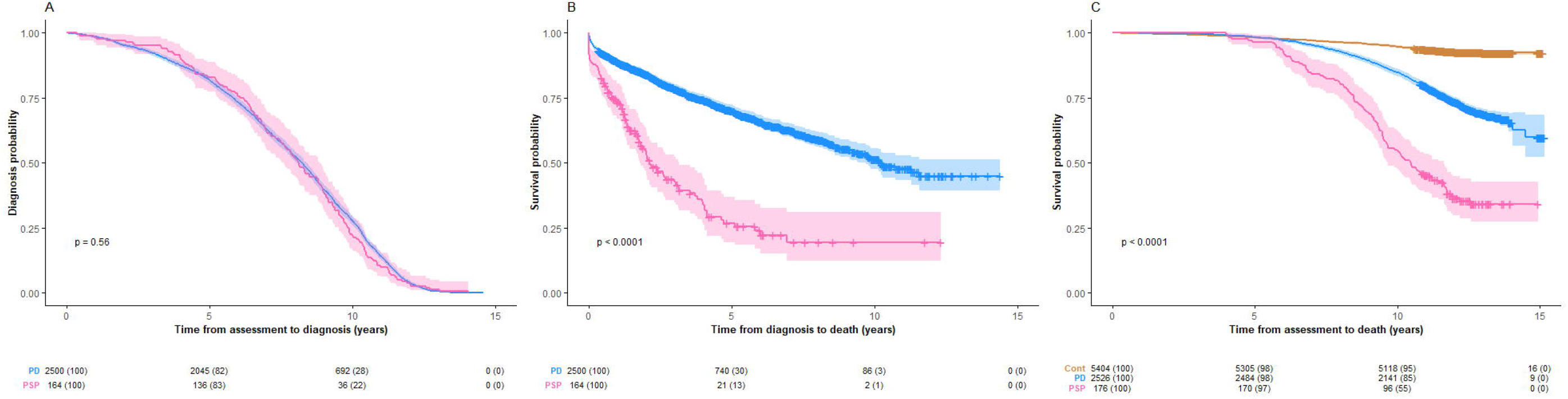
Survival analysis using a Cox Regression model from (A) assessment to diagnosis (B) diagnosis to death and (C) assessment to death split according to diagnostic groups with associated number at risk tables below each plot. Cont = Controls, PD = Parkinson’s disease, PSP = Progressive Supranuclear Palsy.

Baseline quantitative physical and cognitive data are displayed in Table 1 and Figure 2. Apart from fluid intelligence, which was lower in the PSP group, no significant differences were observed between PSP and Parkinson’s disease groups. Apart from a pair matching task, both disease groups were significantly different from controls in all parameters with slower reaction times, weaker handgrip strength and poorer performance on tasks of fluid intelligence and digit recall. Baseline categorical data are displayed in Table 2 and Figure 2. Both PSP and Parkinson’s disease groups reported significantly more falls than the control group. The PSP group reported significantly more falls than the Parkinson’s disease group. Self-rated health score did not differ between the PSP group and the Parkinson’s disease group but both groups reported worse health ratings than the control group. Both the PSP and Parkinson’s disease groups had poorer performance on the task of prospective memory with fewer correct first attempts. The PSP group had more correct second attempts than the Parkinson’s disease group and the control group.

**Table 2:** Categorical data from baseline assessment

|  | Falls in the last year |  |  | Self-rated health score |  |  | Prospective memory |  |  |
| --- | --- | --- | --- | --- | --- | --- | --- | --- | --- |
|  | None<br>(n = 6337) | One or<br>more<br>(n = 1754) | *p-value* | Excellent<br>and Good<br>(n = 6027) | Fair and<br>Poor<br>(n = 2028) | *p-value* | Correct at<br>first attempt<br>(n = 2120) | Incorrect at<br>first attempt<br>(n = 688) | *p-value* |
| **Demographics** |  |  |  |  |  |  |  |  |  |
| Condition: |  |  |  |  |  |  |  |  |  |
| Control (number, %) | 4311 (80) | 1087 (20) | <.001 | 4278 (79) | 1103 (21) | <.001 | 1555 (78) | 443 (22) | <.001 |
| PD (number, %) | 1910 (76) | 608 (24) | - | 1637 (65) | 864 (35) | - | 525 (71) | 219 (29) | - |
| PSP (number, %) | 116 (66) | 59 (34) | - | 112 (65) | 61 (35) | - | 40 (61) | 26 (39) | - |
| Age at baseline (years) | 63.6 (5.4) | 64.1 (5.0) | 0.004 | 63.7 (5.2) | 63.7 (5.5) | .157 | 63.5 (5.2) | 65.0 (4.8) | <.001 |
| Sex, Male/Female (%) | 4077/2260<br>(64/36) | 931/823<br>(53/47) | <.001 | 3660/2367<br>(61/39) | 1322/706<br>(65/35) | <.001 | 1350/770<br>(64/36) | 401/287<br>(58/42) | .013 |
| Age at diagnosis (years) | 71.6 (6.0) | 71.1 (5.9) | .016 | 71.8 (5.9) | 71.1 (6.1) | .006 | 70.6 (6.0) | 71.7 (5.4) | .007 |
| Time to diagnosis (years) | 7.9 (2.9) | 7.5 (3.1) | .007 | 8.0 (2.8) | 7.5 (3.1) | .002 | 7.4 (2.6) | 7.2 (2.8) | .501 |
| **Mortality:** |  |  |  |  |  |  |  |  |  |
| Number dead (%) | 885 (14) | 318 (18) | <.001 | 720 (12) | 476 (24) | <.001 | 237 (11) | 116 (17) | <.001 |
| Age at death (years, SD) | 74.1 (5.0) | 73.7 (5.0) | .148 | 74.3 (5.0) | 73.7 (5.0) | .014 | 72.8 (5.1) | 73.6 (4.5) | .169 |
| Assessment to death (years, SD) | 8.7 (2.8) | 8.4 (2.8) | .045 | 8.9 (2.5) | 8.3 (3.0) | .001 | 8.0 (2.3) | 7.5 (2.7) | .226 |
| Diagnosis to death (years, SD) | 2.3 (2.4) | 2.4 (2.4) | .213 | 2.4 (2.4) | 2.2 (2.4) | .405 | 2.1 (2.2) | 2.0 (2.2) | .525 |
| **Quantitative test parameters:** |  |  |  |  |  |  |  |  |  |
| Weight (kg) | 79.3 (14.6) | 79.5 (15.7) | .856 | 77.7 (13.9) | 84.2 (16.2) | <.001 | 79.9 (15.0) | 78.8 (14.8) | .158 |
| Reaction time (ms) | 578 (119) | 598 (137) | <.001 | 574 (116) | 604 (143) | <.001 | 577 (116) | 624 (144) | <.001 |
| Left hand grip strength (kg) | 31.1 (10.8) | 27.3 (11.4) | <.001 | 30.9 (10.8) | 28.5 (11.6) | <.001 | 30.1 (10.5) | 27.4 (10.7) | <.001 |
| Right hand grip strength (kg) | 33.2 (10.9) | 29.2 (11.2) | <.001 | 33.0 (10.9) | 30.6 (11.4) | <.001 | 32.3 (10.6) | 29.5 (10.7) | <.001 |
| Pair matching (errors) | 4.4 (3.6) | 4.5 (3.5) | .134 | 4.4 (3.6) | 4.6 (3.7) | .166 | 4.0 (3.2) | 5.3 (4.0) | <.001 |
| Fluid intelligence score | 6.1 (2.1) | 5.8 (2.1) | .038 | 6.2 (2.1) | 5.6 (2.2) | <.001 | 6.4 (2.0) | 4.9 (2.0) | <.001 |
| Digit recall score | 6.6 (1.7) | 6.2 (2.0) | .012 | 6.6 (1.7) | 6.0 (2.1) | <.001 | 6.8 (1.5) | 5.8 (2.2) | <.001 |

PD = Parkinson's disease, PSP = Progressive Supranuclear Palsy, SD = standard deviation

**Figure 2:**
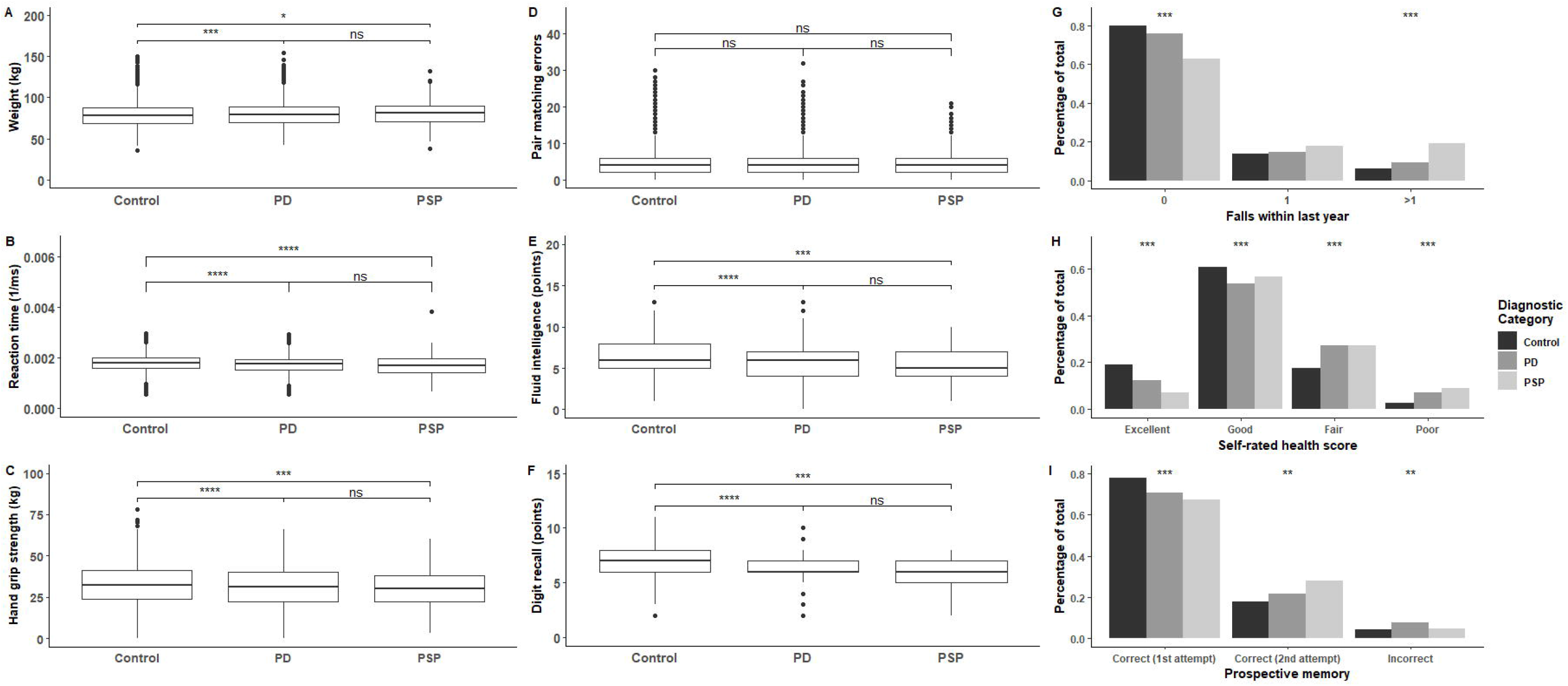
Baseline assessment characteristics by diagnostic group. PD = Parkinson’s Disease, PSP = Progressive Supranuclear Palsy. Adjusted p-value: ns: p>0.05, *: p<=0.05, **: p<=0.01, ***: p<=0.001, ****: p<=0.0001

Categorical data collected at baseline assessment and combined across disease groups are also displayed in Table 2. Individuals reporting more falls were older at baseline but younger at diagnosis with a shorter time to diagnosis, a higher mortality and a shorter time from assessment to death. More falls also correlated with slower reaction time, weaker handgrip and female sex. Worse subjective self-rated health scores at baseline correlated with male sex, a younger age at diagnosis, a shorter time to diagnosis, a higher mortality and a shorter time from assessment to death. Increased weight, slower reaction times, weaker handgrip strength and poorer cognitive performance correlated with “Fair” or “Poor” health scores. Poorer performance on the prospective memory task correlated with poorer performance on other physical and cognitive parameters, a higher mortality, female sex and an older age at baseline.

Kaplan-Meier curves for both disease groups from baseline assessment to diagnosis and diagnosis to death, and all groups from baseline assessment to death, are presented in Figure 1. No baseline assessment parameters predicted a shorter survival in the PSP group. With relation to baseline assessments, a shorter time to diagnosis was associated with male sex (HR 1.45, 95% CI, 1.04-2.03, p=.029) in the PSP group. A self-rated health score of “Poor” (HR 1.25, 95% CI 1.03-1.51, p=0.024) was associated with a shorter time to diagnosis in the Parkinson’s disease group and when combining both disease groups (HR 1.26, 95% CI, 1.05-1.52, p=.014). Age (HR 1.02, 95% CI 1.01-1.03, p<0.001) and self-rated health scores of “Good” (HR 1.20, 95% CI 1.06-1.36, p=0.005) and “Fair” (HR 1.31, 95% CI 1.14-1.50, p<0.001) were associated with poorer survival in the Parkinson’s disease group.

## Discussion

The principal results of this study are that people who will later develop PSP differ from those who do not, as early as an average of seven years before diagnosis. They manifest differences in reaction time, hand grip strength, fluid intelligence, prospective memory, self-rated health score and digit recall tasks, and a tendency to falls. These prodromal features are similar to those previously described in Parkinson’s disease.^21,22^ We suggest that these features are present approximately 4 years before symptom onset given the average three years’ interval between first symptom and PSP diagnosis in multiple observational studies.^2,6,23^ We also confirm the significantly faster progression and poorer survival in those developing PSP compared to Parkinson’s disease.

Changes in those later developing Parkinson’s disease were similar to previous studies.^21,22^ Subtle motor dysfunction, measurable as slowed reaction time and decreased hand grip strength bilaterally, was present over seven years prior to diagnosis. Performance on tasks of fluid intelligence and digit recall was poorer than age and gender matched controls. Older age and worse self-rated health scores were associated with shorter survival. Moreover, lower self-rated health scores were more frequent than controls and had strong predictive values of poorer performance on tasks of physical and cognitive functioning, supporting the utility of this candidate marker described in the most recent diagnostic criteria.^9^

Apart from poorer fluid intelligence scores in the pre-PSP group, no other differences in quantitative data were detected between the pre-Parkinson’s disease and pre-PSP groups at study baseline. Almost a third of the PSP cohort were initially diagnosed with Parkinson’s disease, indicating that a high degree of initial diagnostic uncertainty or error exists. As a group, pre-PSP patients had more falls in the year prior to baseline assessment than both pre-Parkinson’s disease patients and age and gender-matched controls, but falls were only reported by a minority (29/176, 16%). Mortality findings were similar to previous studies^6,21,24,25^ with PSP demonstrating shorter survival, younger age at death, shorter time from assessment to death, and shorter time from diagnosis to death compared to Parkinson’s disease.

An advantage of this study is the large sample size incorporating both primary and secondary care data which provides a truer representation of the prodromal period that is not reliant on recall or selection bias. However, there are several limitations. Firstly, the baseline tests measured are neither specific nor sensitive for the detection of Parkinson’s disease or PSP. The physical tests do not include any proxy markers for rigidity, tremor or eye movement abnormalities and the cognitive tests no assessment of executive function or fluency; an important distinguishing feature between early PSP and Parkinson’s disease.^26^ There are also no screening questions for related highly predictive prodromal features and conditions (e.g. anosmia, REM-sleep behaviour disorder). Secondly, the accuracy of diagnosis is reliant on a ‘real-world’ setting; neither research criteria fulfilment nor level of movement disorder expertise can be guaranteed. Thirdly, the UKB cohort is not representative of the UK population with regards to race, ethnicity and socioeconomic factors that may be relevant to both Parkinson’s disease and PSP risks, and their prognosis. Fourthly, there is no pathological confirmation nor record made of treatment or responsivity to levodopa medications.

In contrast to Parkinson’s disease, comparatively little is known about the pre-symptomatic and prodromal periods in PSP. Once the diagnosis is confirmed, the disease is often at an advanced stage with prognosis universally poor. Our data show deterioration in simple physical and cognitive markers detectable over seven years before PSP diagnosis. These changes were similar to those detected in the Parkinson’s disease group and support the hypothesis of a long prodromal phase in PSP. The differences in cognitive markers in particular would benefit from deeper phenotyping. The results from the UK Biobank cohort will help to inform early diagnosis and stratification of PSP for future disease modifying trials.^27,28^

## Data Availability

UK Biobank data are available through a procedure described at http://www.ukbiobank.ac.uk/using-the-resource/.

## Acknowledgement

This research has been conducted using data from the UK Biobank, a major biomedical database. The UK Biobank is generously supported by its founding funders the Wellcome Trust and UK Medical Research Council, as well as the Department of Health, Scottish Government, the Northwest Regional Development Agency, British Heart Foundation and Cancer Research UK. Our study was funded by the National Institute for Health Research (NIHR) Biomedical Research Centre and Biomedical Research Unit in Dementia based at Cambridge University Hospitals NHS Foundation Trust and the University of Cambridge (BRC-1215-20014); the Cambridge Centre for Parkinson-plus (RG95450); the Wellcome Trust (103838/Z/14/Z); and the Medical Research Council (MR/P01271X/1; SUAG051/G101400). The views expressed are those of the authors and not necessarily those of the NIHR or the Department of Health and Social Care.

## Authors’ Roles

D Street: 1A, 1B, 1C, 2A, 2B, 2C, 3A, 3B

D Whiteside: 2A, 2B, 2C, 3B

T Rittman: 2C, 3B

JB Rowe: 1A, 1B, 1C, 2A, 2C, 3A, 3B

